# The Aortic Flow Reversal Ratio: A Quantitative Adjunct to the Bicêtre Score in Vein of Galen Malformation

**DOI:** 10.1101/2025.08.01.25332767

**Authors:** Menachem Rimler, Ranjit Philip, Lydia Henry, Hannah Huth, Lucas Elijovich

## Abstract

**Objective:** The Bicêtre score is a clinical tool used to guide the timing of intervention in Vein of Galen Aneurysmal Malformation (VGAM). However, it relies on the presence of end-organ damage. This study aimed to determine if an echocardiographic parameter, the Aortic Flow Reversal Ratio (AoFRr), can quantify significant systemic steal in clinically stable neonates (Bicêtre score ≥12) and to evaluate its utility in predicting treatment outcomes.

**Methods:** We conducted a single-center retrospective study in patients with VGAM who underwent endovascular embolizations. Transthoracic echocardiography was used to calculate the AoFRr (diastolic reversal VTI / systolic forward VTI) before and after intervention. We analyzed the prevalence and degree of pre-intervention flow reversal. Linear regression was used to correlate the pre-intervention AoFRr with the Bicêtre score. The change in AoFRr post-intervention was evaluated for its association with the likelihood of requiring subsequent embolizations using Wilcoxon signed-rank and Chi-square tests.

**Results:** In the cohort of 12 patients with a median total Bicêtre score of 18 (IQR 15.5 - 20), 83.3% demonstrated pre-intervention aortic diastolic flow reversal. The median pre-intervention AoFRr was 0.81 (IQR 0.49 - 1.05), indicating substantial systemic steal. Pre-intervention AoFRr moderately correlated with the Bicêtre score (R² = 0.4546). The initial embolization resulted in a statistically significant mean decrease in the AoFRr of 52.80% (p = 0.0232). A post-intervention reduction in AoFRr of ≥85% was significantly associated with a lower likelihood of requiring re-intervention (p = 0.0253).

**Conclusion:** Significant hemodynamic steal, quantified by the AoFRr, is evident on echocardiography in VGAM patients even when they are considered clinically stable by the Bicêtre score. The AoFRr provides a valuable, non-invasive measure of hemodynamic compromise that correlates with clinical severity scores. Its reduction following embolization predicts a more favorable clinical course. The AoFRr may serve as a critical adjunct to the Bicêtre score for risk stratification and for optimizing the timing of endovascular intervention.

## Introduction

Vein of Galen Aneurysmal Malformation (VGAM) is a rare congenital intracranial arteriovenous malformation that shunts blood from arterial feeders to the persistent median prosencephalic vein of Markowski, a precursor to the vein of Galen^1–5^. This low-resistance shunt (Figure 1B) steals blood from the systemic circulation, a phenomenon known as systemic steal. This “steal” redirects cardiac output away from the body’s organs and toward the fistula, causing diastolic reversal of flow in the aorta (Figure 1D) and leading to significant hemodynamic disturbances^5–7^. Due to the redirection of cardiac output through the shunt, there is reduced perfusion of vital organs. The high velocity shunt bypasses normal gas exchange in the brain^8,9^. The consequent high venous return from the shunt causes right heart volume overload and high-output cardiac failure, which can be life-threatening in neonates^1,10–13^ (Figure 1).

**Figure 1.**
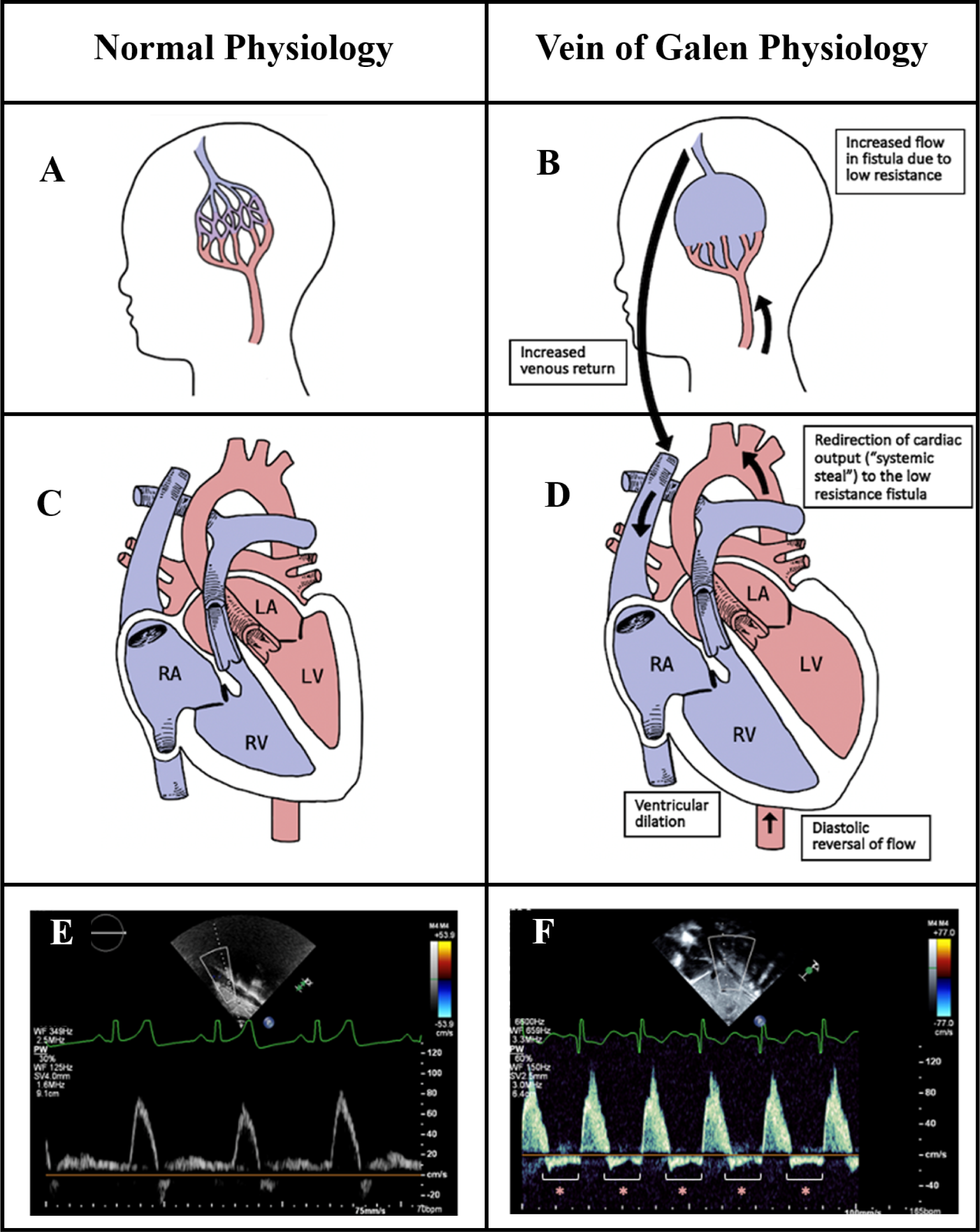
Pathophysiology of Vein of Galen Aneurysmal Malformation. (A) Normal cerebral arterial and venous anatomy. (B) In VGAM, a low-resistance fistula shunts arterial blood directly into a persistent embryonic vein, leading to increased flow and venous return. (C) Normal cardiac physiology with balanced systemic circulation. (D) VGAM physiology demonstrates the consequences of the shunt, including redirection of cardiac output (“systemic steal”) toward the fistula, causing diastolic reversal of flow in the aorta and subsequent right-sided heart volume overload and ventricular dilation. (E) A normal Doppler flow pattern in the abdominal aorta shows only forward (anterograde) flow. (F) In VGAM, the Doppler pattern reveals significant diastolic flow reversal (retrograde flow as indicated by red asterisks), indicative of aortic steal.

The hemodynamic effects can lead to respiratory distress, heart failure, and neurological abnormalities^14,15^. Without intervention, this condition can lead to multi-organ failure and death^1,2,16^. Endovascular embolization is the primary treatment modality aimed at reducing blood flow through the shunt^17,18^. However, the timing of intervention is critical, as both premature and delayed intervention can lead to complications^19–21^.

The Bicêtre score is a clinical tool that evaluates multiple parameters (cardiac, respiratory, neurological, hepatic and renal function) and has traditionally been used to guide the neuro-interventionalist on optimal timing of the procedure^4,22^. It categorizes patients based on severity, with scores less than 8 indicating a near-fatal prognosis and futility of intervention, scores between 8 and 12 suggesting benefit from emergent endovascular embolization (EE), and scores greater than 12 indicating candidates for medical management until around 5 months of age. It is used as a proxy for grading end organ damage^11,23–25^.

Transthoracic echocardiograms (TTE) are routinely performed in neonates with VGAM to assess the hemodynamic changes resulting from the arteriovenous shunt, including pulmonary hypertension and high-output heart failure. It provides critical information on the severity of the condition and response to medical management^1,26–28^.

In these patients, systemic steal can be visualized on TTE as diastolic flow reversal in the aorta (Figure 1F), a stark contrast to the forward-only flow seen in normal physiology (Figure 1E)^29,30^. This reversal can be quantified using the Aortic Flow Reversal Ratio (AoFRr), which is calculated as the ratio of the diastolic reversal velocity time integral (VTI) to the systolic forward VTI (Figure 2). While the aortic flow reversal is a known surrogate for systemic steal in other conditions with shunts, such as a patent ductus arteriosus (PDA), its specific utility in VGAM remains poorly defined^31,32^. This represents a critical gap, as management decisions currently rely on clinical scores that reflect existing end-organ damage rather than the underlying hemodynamic stress. This study, therefore, aims to determine if the AoFRr can quantify systemic steal in clinically stable neonates with VGAM, providing a potential adjunct to the Bicêtre score for optimizing intervention timing before irreversible organ damage occurs.The hypothesis of this study is that TTE can be used to estimate significant systemic steal even in the absence of end organ damage (as estimated by the Bicêtre score).

**Figure 2.**
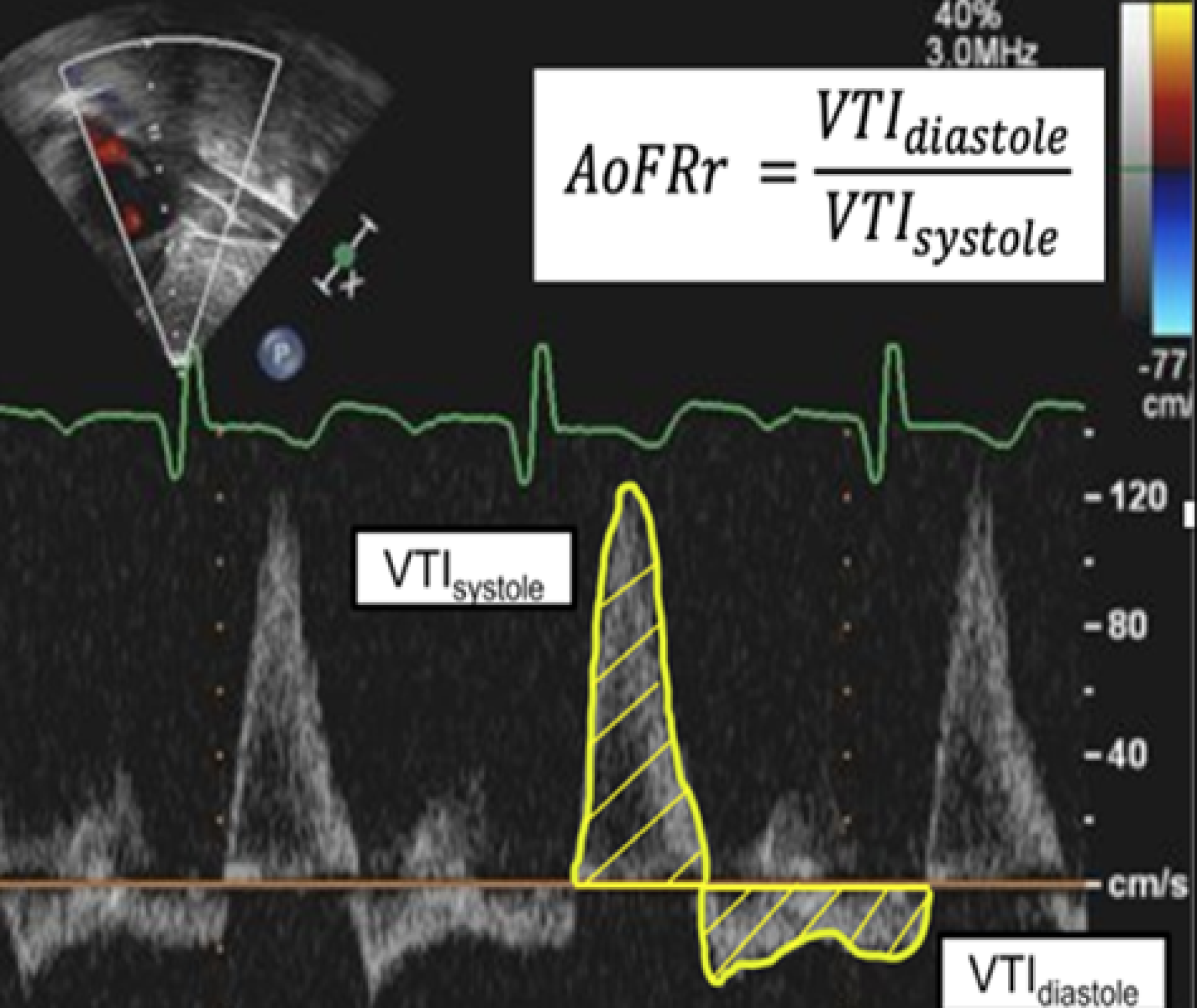
Echocardiographic Quantification of the Aortic Flow Reversal Ratio (AoFRr). A pulse-wave Doppler tracing from the abdominal aorta is used to calculate the AoFRr. The Velocity Time Integral (VTI), which is proportional to stroke volume, is measured by tracing the Doppler waveform. The systolic VTI (VTI_systole_) represents the anterograde flow during systole (waveform above the baseline), while the diastolic reversal VTI (VTI_diastole_) represents the retrograde flow stolen by the malformation during diastole (waveform below the baseline). The AoFRr is calculated as the ratio of these two values.

The primary outcome is to evaluate the pre-intervention flow reversal by TTE in patients with high Bicêtre scores. The secondary outcomes were 1) to correlate the Bicêtre score to the degree of reversal of flow, 2) to assess the hemodynamic impact of the intervention based on the change in the TTE derived AoFRr by comparing the degree of change in AoFRr to the likelihood of undergoing additional EE.

## Methods

### Study Design

We conducted a retrospective cohort study of patients treated at the University of Tennessee Health Science Center, Le Bonheur Children’s Hospital, Memphis, TN, over a 10-year period. This study was approved by the Institutional Review Board (IRB) at Le Bonheur Children’s Hospital (IRB # 24-10378-XM).

### Patient Selection

We identified patients through a systematic review of institutional electronic medical records and radiology databases and verified the diagnosis of VGAM according to established diagnostic guidelines. To be included in this study, patients were required to meet the following criteria:

1. A confirmed diagnosis of VGAM based on imaging evidence.
2. A documented Bicêtre Neonatal Evaluation Score of greater than 12 at the time of initial assessment.
3. At least one EE procedure performed during the study period.
4. Availability of complete pre-and post-intervention TTE datasets.

We excluded patients with incomplete echocardiographic data or an unconfirmed diagnosis. The final selection process involved cross-referencing clinical notes with procedural records to confirm eligibility, ensuring that only infants with comprehensive and relevant data were included.

## Data Collection

We organized data collection into two main categories, clinical and echocardiographic, each requiring specific methods to ensure accuracy and reliability.

Clinical Data: We evaluated the clinical condition before intervention using Bicêtre scores, a standardized system that measures multi-organ function (including cardiac, cerebral, respiratory, hepatic, and renal systems) in patients with VGAM. Clinicians extracted these scores from electronic medical records using a predefined procedure. We also recorded the patient’s age at intervention and the total number of EE procedures. The latter was used in our secondary analysis as a measure of treatment frequency and complexity.

Echocardiographic Data: A board-certified pediatric cardiologist reviewed TTE images obtained before and after each EE procedure for consistency. The primary focus was the AoFRr, a measurement used to assess the hemodynamic impact of VGAM. Doppler ultrasound of the abdominal aorta provides VTI measurements for both diastolic flow reversal (retrograde flow during diastole) and forward systolic flow (anterograde flow during systole). We calculated the diastolic reversal VTI by manually tracing the retrograde Doppler waveform and the systolic forward VTI by tracing the anterograde waveform. We then calculated the AoFRr as the ratio of the diastolic reversal VTI to the systolic forward VTI, expressed as a unitless value that correlates with the relative amount of shunted blood flow through the VGAM, as supported by prior studies of arteriovenous shunting^11,27,33–35^ (Figure 2).

## Statistical Analysis

We performed all statistical analyses using SPSS Statistics v21.0 (IBM Corp, Armonk, New York), with statistical significance set at a p-value of less than 0.05.

We used descriptive statistics to characterize the study cohort. Continuous variables, such as the pre-intervention AoFRr, are presented as medians and interquartile ranges (IQR). Categorical data, including the presence or absence of flow reversal, are reported as percentages. We determined the statistical significance of the percentage decrease in AoFRr following the initial endovascular procedure using a Wilcoxon signed-rank test for paired samples.

To evaluate the secondary outcomes, we conducted the following analyses:

1. A simple linear regression to assess the relationship between the pre-intervention AoFRr and the Bicêtre score, with the coefficient of determination (R²) reported to quantify the correlation.
2. An assessment of the association between the reduction in flow and the number of subsequent interventions using two methods. First, we calculated a Spearman’s rank correlation to measure the relationship between the ranked absolute flow reduction (AFR) and the total number of re-interventions. Second, we dichotomized the percentage drop in AoFRr using an 85% reduction as a cutoff and used a Chi-square test of independence to determine its association with the likelihood of requiring any re-intervention.

## Results

### Patient Demographics and Clinical Characteristics

A total of 12 patients met the inclusion criteria for this study. The cohort was predominantly male (n=7, 58.3%). The median age at the time of the initial endovascular intervention was 3.7 months (IQR 0.3 - 39.5 months), with a median weight of 3.4 kg (IQR 3.0 - 6.2 kg) and a median height of 51.5 cm (IQR 50.0 - 54.0 cm). Over the study period, the cohort underwent a total of 30 endovascular interventions and 49 TTEs. The median number of interventions per patient was 2 (IQR 1 - 3), and the median number of echocardiograms per patient was 5 (IQR 3 - 8). All interventions were performed via a transfemoral arterial route using liquid embolic embolization of the VOGM fistulous connections with n-butyl cyanoacrylate. As per the study’s inclusion criteria, all patients had a pre-intervention Bicêtre Neonatal Evaluation Score greater than 12.

The median score for the cohort was 18 (IQR 15.5 - 20). The median scores for the clinical subcategories were as follows: Cardiac 5 (IQR 2.5 - 5), Cerebral 5 (IQR 5 - 5), Respiratory 5 (IQR 2.5 - 5), Hepatic 3 (IQR 3 - 3), and Renal 3 (IQR 3 - 3).

### Baseline Echocardiogram Findings and Correlation with Bicêtre scores

Prior to the initial endovascular intervention, diastolic flow reversal in the abdominal aorta was a common finding, present in 83.3% of the patients. The median pre-intervention AoFRr for the cohort was 0.81 (IQR 0.49-1.05), indicating a significant degree of systemic steal even in patients with high Bicêtre scores. A simple linear regression analysis demonstrated a moderate correlation between the pre-intervention AoFRr and the total Bicêtre score, yielding an R-squared value of 0.4546.

### Hemodynamic Impact of Endovascular Intervention

The initial EE procedure had a significant hemodynamic impact, resulting in a mean percentage decrease in the AoFRr of 52.80% (p = 0.0232). The degree of flow reduction after the first intervention was associated with the need for subsequent procedures. A Spearman’s rank correlation showed a moderate negative relationship between the absolute flow reduction (AFR) and the number of re-interventions (ρ ≈-0.55, where ρ represents the Spearman’s rank correlation coefficient). Furthermore, when the percentage drop in AoFRr was dichotomized at a cutoff of 85%, a Chi-square test revealed that patients achieving this level of reduction were significantly less likely to require any re-intervention (χ² = 5, p = 0.0253).

## Discussion

Infants with VGAM present significant challenges for clinicians in prognostication and management. The Bicêtre score is a valuable clinical tool used to assess the severity of heart failure and multi-organ dysfunction. This study focusses on the adjunct use of TTE as a non-invasive metric to evaluate the hemodynamic significance of VGAM in providing an insight into not only the timing of intervention but also the hemodynamic effect of the intervention. TTE is an essential, non-invasive tool in the initial diagnosis and ongoing management of neonates with VGAM. Its primary role has been to evaluate the profound hemodynamic consequences of the high-flow intracranial shunt, most notably high-output cardiac failure^27^. Previous studies have consistently identified hallmark structural findings on TTE, including dilation of the right atrium and ventricle, flattening of the interventricular septum, and a dilated superior vena cava due to the massive venous return from the malformation ^36–38^(Figure 1B and 1D).Beyond structural assessment, Doppler ultrasonography is critical for identifying prognostic markers^11,27,34,39^. The phenomenon of diastolic flow reversal in the descending aorta, or “aortic steal,” is a well-documented qualitative marker of a significant shunt^31–33,40,41^ (Figure 1F). To better quantify this, several echocardiographic methods have been described. Schwarz et al. proposed a “percentage of steal” using the time-averaged maximum velocity (TAMAX) to represent retrograde flow as a proportion of total flow (Retrograde TAMAX) / (Antegrade TAMAX + Retrograde TAMAX)^34^. In contrast, others have used the VTI, a robust and reproducible measure of flow distance proportional to volume. Doctor et al. used VTI to create an antegrade-to-retrograde ratio (Antegrade VTI / Retrograde VTI), identifying a ratio <1.5 as predictive of mortality^27^. In our study, we also utilized VTI but calculated the Aortic Flow Reversal Ratio as the inverse (Retrograde VTI / Antegrade VTI) (Figure 2). We chose this method because expressing the abnormal retrograde flow as the numerator provides a more clinically intuitive value; a higher number directly corresponds to greater systemic steal, with a ratio approaching 1.0 representing a near-equal backward and forward flow volume. This approach grounds our metric in a standard echocardiographic technique that is both physiologically representative of volume steal and easily replicable. By manually tracing the Doppler waveform, VTI captures the integral of velocity over time, a value directly proportional to stroke volume^42–44^. This makes it a more accurate reflection of the true volume of blood being shunted away from the systemic circulation compared to methods based on instantaneous or averaged velocities. Furthermore, because VTI tracing is a fundamental skill in sonography and a common function on modern ultrasound systems, this metric can be readily adopted and consistently measured across different institutions without requiring specialized software or complex calculations. While a direct mathematical conversion between our VTI-based AoFRr and the TAMAX-based’percentage of steal’ used by Schwarz et al. is not possible due to the different underlying measurements, both ratios serve the same fundamental purpose: to quantify the proportion of retrograde flow relative to antegrade flow as a surrogate for the severity of the systemic steal.

The principle of using aortic diastolic flow reversal as a surrogate for systemic steal is not unique to VGAM and is a well-established concept in the assessment of other hemodynamically significant shunts, such as a PDA^32,35,45^. In the presence of a moderate to large PDA with low pulmonary vascular resistance, one of the expected findings is holo-diastolic flow reversal at the descending aorta^31,33^. Reinforcing its value in assessment of hemodynamic significance, a study by Broadhouses et al. investigating various echo markers in comparison to cardiac MRI, found that diastolic flow reversal in the descending aorta had the best correlation with a hemodynamically significant PDA when compared to other parameters^45^.

The primary finding of this investigation is the profound disconnect between clinical appearance and underlying hemodynamic reality in infants with VGAM. We found that significant systemic steal, as quantified by the AoFRr, is highly prevalent even in a cohort considered clinically stable with a Bicêtre score of 12 or greater. While the Bicêtre score remains an invaluable tool for assessing overt, end-stage multi-organ dysfunction, our results suggest its shortcoming as an early diagnostic tool for hemodynamic significance. An overwhelming majority of these “stable” patients (83.3%) exhibited flow reversal with a median pre-intervention AoFRr of 0.81, indicating that 81% of the blood pumped forward by the heart was stolen back into the low-resistance shunt. For context, the mortality-predictive threshold of <1.5 reported by Doctor et al. is mathematically equivalent to an AoFRr >0.67 in our metric^27^. The fact that our cohort’s median value so clearly surpasses this high-risk hemodynamic threshold— all while maintaining reassuring clinical Bicêtre scores—is a striking and critical observation.

This highlights a critical insight: a reassuring Bicêtre score can mask a state of significant, ongoing hemodynamic stress. The AoFRr provides a direct, non-invasive measure of this systemic steal, offering a quantitative window into the underlying pathophysiology. It allows clinicians to see the fraction of cardiac output being diverted from the body long before this chronic burden translates into the overt clinical end-organ damage required to lower the Bicêtre score. This finding directly addresses a key gap in risk stratification, demonstrating that the AoFRr can unmask high-risk hemodynamic compromise in a subclinical phase. It therefore has the potential to serve as a more sensitive, early indicator to guide the timing of intervention, suggesting that endovascular embolization could be beneficial in mitigating this silent burden even in patients who appear clinically well.

Perhaps the most clinically actionable finding of this study is the association between the post-procedural reduction in AoFRr and the need for future interventions. The initial endovascular embolization resulted in a statistically significant mean decrease in the AoFRr of 52.80% (p = 0.0232). This intervention effectively reduced the cohort’s median AoFRr from a high-risk value of 0.81 to approximately 0.38—a level well below the critical threshold of 0.67 derived from the Doctor et al. data^27^. This is not merely a statistical change; it represents a clear and quantifiable hemodynamic improvement, reflecting a dramatic reduction in the steal phenomenon and a restoration of more effective systemic perfusion. This immediate, measurable impact on aortic flow dynamics is what gives the metric its profound clinical utility. The magnitude of this improvement has a dual utility:

1. **Determining Procedural Success:** For the interventionalist, the change in AoFRr offers immediate, objective feedback on the hemodynamic success of an embolization. It redefines procedural success, shifting the focus from simple anatomical occlusion to quantifiable physiological improvement. Achieving a significant reduction in the AoFRr (e.g., ≥85% as identified in our analysis) could serve as a novel therapeutic endpoint for the initial procedure.
2. **Prognostication and Family Counseling:** For both the clinical team and the patient’s family, the magnitude of the post-procedural drop in AoFRr is a powerful prognostic tool. It allows for more informed counseling regarding the expected clinical course. A large reduction can provide reassurance and potentially suggest a lower likelihood of requiring further interventions, whereas a minimal reduction can help set expectations that despite a technically successful procedure, further EE will likely be necessary.

## Limitations

This study’s findings are constrained by several factors. Its retrospective, single-center design and a small cohort limit statistical power and generalizability. Furthermore, a significant selection bias was introduced by including only patients with a Bicêtre score greater than 12.

This focus was necessary as a review of institutional records revealed an insufficient number of patients with a score of 12 or less to perform a meaningful comparative analysis. Consequently, our conclusions regarding the AoFRr cannot be extrapolated to the entire VGAM population, particularly the most critically ill infants, and require validation in larger, multi-center prospective studies. Additionally, our secondary outcomes focused on a procedural endpoint (need for re-intervention) rather than long-term clinical data. Our analysis, therefore, does not establish a direct link between AoFRr and ultimate neurodevelopmental outcomes, which remains a the most critical outcome which requires future investigation.

### Future Directions

While this study provides valuable preliminary evidence for the utility of the AoFRr in a select population of infants with VGAM, several avenues for future research are necessary to validate and expand upon these findings.

First, a large-scale, multi-center prospective study is the critical next step. Such a study would overcome the limitations of our small, single-center cohort, allowing for greater statistical power and the generalizability of our results. A prospective design would enable standardized protocols for both echocardiographic measurement and clinical data collection, reducing potential biases inherent in retrospective analyses.

This future research should aim to enroll patients across the entire clinical spectrum of VGAM, including those presenting with severe heart failure and low Bicêtre scores. By analyzing the AoFRr in a more heterogeneous population, a more robust risk-stratification model could be developed, potentially identifying different hemodynamic thresholds for intervention based on the initial clinical severity and VOGM subtype (choroidal vs. mural).

Furthermore, a crucial area for subsequent investigation is the correlation between the AoFRr and long-term neurodevelopmental outcomes. Our study focused on the procedural endpoint of re-intervention; however, the ultimate goal of treatment is to ensure the best possible neurological and cognitive function. Longitudinal studies that track patients from infancy through childhood are needed to determine if early correction of the hemodynamic steal, as measured by the AoFRr, translates into improved long-term outcomes.

Ultimately, the findings from this and subsequent observational studies could provide the foundation for a prospective interventional trial. Such a trial would not seek to replace the validated, multi-system Bicêtre score, but rather to determine if a new treatment algorithm that integrates the AoFRr as an adjunct to clinical assessment could lead to improved outcomes. This approach could help refine the management paradigm by identifying a window for proactive intervention based on quantifiable hemodynamic data, potentially before the clinical decline that lowers the Bicêtre score. This line of inquiry could lead to more personalized and timely interventions, with the goal of improving the overall prognosis for this vulnerable patient population.

## Conclusion

The AoFRr is a valuable, non-invasive metric that provides a real-time, quantifiable assessment of the primary pathophysiologic insult in VGAM. It can unmask severe hemodynamic derangement in infants who may otherwise appear clinically stable. Used as an adjunct to the Bicêtre score, the AoFRr has the potential to substantially refine clinical management by enabling a more proactive treatment strategy. It provides precision medicine by guiding the timing of intervention based on hemodynamic severity rather than waiting for signs of end-organ failure. The magnitude of AoFRr reduction of 85% or more following the initial EE is a valuable predictor of a lower likelihood of requiring subsequent procedures, thus providing a measurable target to define procedural success. Ultimately, integrating the AoFRr into routine assessment could shift the paradigm from reactive treatment of organ failure to proactive, hemodynamically-guided management, potentially improving long-term outcomes for this vulnerable population.

## Data Availability

Data will be available upon request

---

| **<u>Demographics</u>** |  |
| --- | --- |
| Age at Initial Intervention (Months) | 3.7 (0.3 - 39.5) |
| Weight at Initial Intervention (Kg) | 3.4 (3.0 - 6.2) |
| Height at Initial Intervention (cm) | 51.5 (50.0 – 54.0) |
| Number of Interventions: total, median (IQR) | 30, 2 (1.8 – 3.0) |
| Number of Echocardiograms: total, median (IQR) | 49, 5 (3 - 8) |
| Male: n (%) | 7 (58.3%) |
| *Data above are Median (Interquartile Range) unless otherwise specified |  |

| **<u>Bicêtre Scores</u>** |  |
| --- | --- |
| **Category** | **Median (IQR)** |
| Cardiac | 5.0 (2.5 – 5.0) |
| Cerebral | 5.0 (5.0 – 5.0) |
| Respiratory | 5.0 (2.5 – 5.0) |
| Hepatic | 3.0 (3.0 – 3.0) |
| Renal | 3.0 (3.0 – 3.0) |
| Total | 18.0 (15.5 – 20.0) |

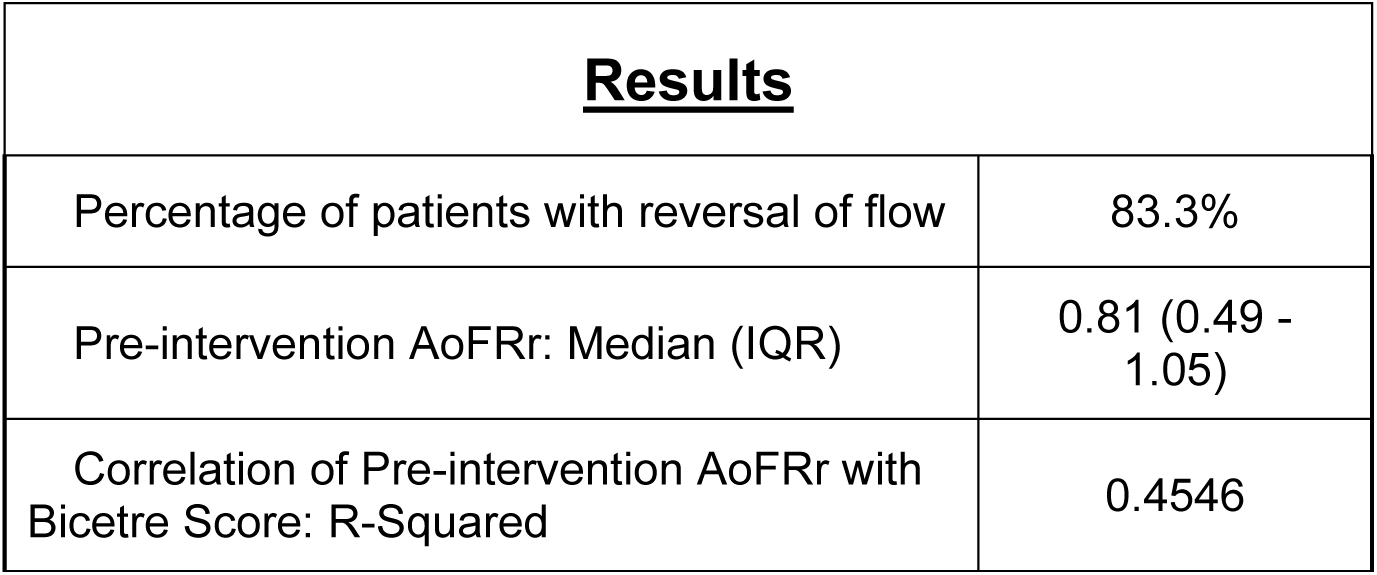
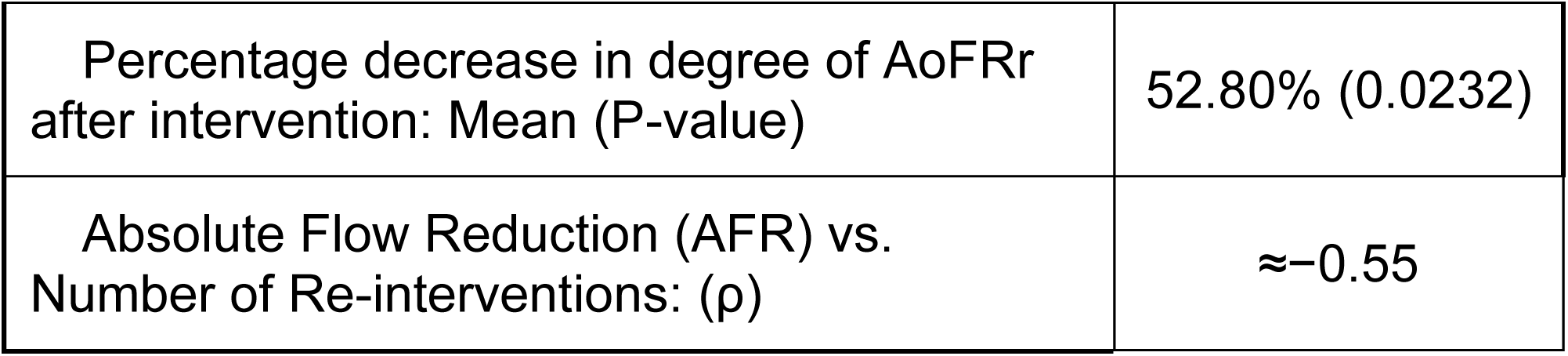

